# Impact of monoclonal antibody treatment on hospitalization and mortality among non-hospitalized adults with SARS-CoV-2 infection

**DOI:** 10.1101/2021.03.25.21254322

**Authors:** J. Ryan Bariola, Erin K. McCreary, Richard J. Wadas, Kevin E. Kip, Oscar C. Marroquin, Tami Minnier, Stephen Koscumb, Kevin Collins, Mark Schmidhofer, Judith A. Shovel, Mary Kay Wisniewski, Colleen Sullivan, Donald M. Yealy, David A Nace, David T. Huang, Ghady Haidar, Tina Khadem, Kelsey Linstrum, Christopher W. Seymour, Stephanie K. Montgomery, Derek C. Angus, Graham M. Snyder

## Abstract

**Background:** Monoclonal antibody (mAb) treatment may prevent complications of COVID-19. We sought to quantify the impact of bamlanivimab monotherapy on hospitalizations and mortality, as well as Emergency Department (ED) visits without hospitalization, among outpatients at high risk of COVID-19 complications.

**Methods:** We compared patients receiving mAb to patients who met criteria but did not receive mAb from December 2020 through March 2021. The study population selection used propensity scores to match 1:1 by likelihood to receive mAb. The primary outcome was hospitalization or all-cause mortality within 28 days; the secondary outcome was hospitalization or ED visit without hospitalization within 28 days. Odds ratios (OR) calculation used logistic regression modeling including propensity score and mAb receipt predictors.

**Results:** The study population included 234 patients receiving mAb and 234 matched comparator patients not receiving mAb. Patients receiving mAb were less likely to experience hospitalization or mortality (OR 0.31, 95% confidence interval [95%CI] 0.17-0.56, p=0.00001) and hospitalization or ED visit without hospitalization (OR 0.50, 95%CI 0.43-0.83, p=0.007). The impact of mAb was more pronounced in prevention of hospitalization (among all age groups, OR 0.35, 95%CI 0.19-0.66, p=0.001) than mortality or ED visit without hospitalization, and most strongly associated with patients age 65 years and older (primary outcome OR 0.28, 95%CI 0.14-0.56, p=0.0003).

**Conclusions:** Bamlanivimab monotherapy was associated with reduction in the composite outcome of hospitalizations and mortality in patients with mild-moderate COVID-19. The benefit may be strongest in preventing hospitalization in patients ages 65 years or older.

## INTRODUCTION

Monoclonal antibodies (mAbs) bind to the SARS-CoV-2 spike (S) protein and block viral entry into host cells, neutralizing the virus.^1-5^ Between November 2020 and February 2021 four mAbs provided as three treatments received Food and Drug Administration Emergency Use Authorization (EUA) for treatment of patients with mild to moderate COVID-19 within 10 days of symptom onset: bamlanivimab 700mg (LY-coV555; Eli Lilly), etesevimab 1,400mg (LY-CoV016; Eli Lilly), casirivimab 1,200mg (REGN10933; Regeneron), imdevimab 1,200mg (REGN10987). Several clinical trials currently evaluate mAbs for prevention or treatment of COVID-19; however, real-world data are limited, and the role of mAbs for patients with COVID-19 remains controversial.^3, 5^

Use of mAb therapy is low in the United States despite widespread drug availability due to lack of robust efficacy data, operational challenges with outpatient infusions, and patient access issues.^6^ Our health system established a mAb program in November 2020 to decrease COVID-19-related complications for patients with mild-moderate illness and expand access to care for underserved patients with COVID-19. Initially, only bamlanivimab monotherapy was available; our evaluation and distribution process has been described elsewhere.^7^ This study quantifies the impact of bamlanivimab monotherapy on hospitalizations, mortality, and Emergency Department (ED) visits among outpatients at high risk of progressing to severe COVID-19. We also explored whether patient age, body mass index, and timing of infusions relative to initial diagnosis had any association with response to therapy.

## METHODS

### Study Setting

UPMC is a 40-hospital integrated academic healthcare system providing care principally within central and western Pennsylvania (USA). After the November 2020 EUA was granted for bamlanivimab, UPMC rapidly established 16 outpatient infusion centers across all served geographical areas. We also infused mAb at our UPMC Senior Communities (i.e., long-term care facilities), patient homes (via collaboration with a home infusion company), and behavioral health units. Physicians referred patients via outpatient and oncology electronic medical records (EMR) or paper order including non-UPMC providers. A centralized team with pharmacists and physicians reviewed orders daily to confirm EUA criteria; decentralized nursing teams then contacted and scheduled eligible patients for infusions. This study was approved by the UPMC Quality Improvement Review Committee (Project ID 2882 and Project ID 3116).

We used the EMR to access all key clinical data, including detailed sociodemographic and medical history data, diagnostic and clinical tests conducted, surgical and other treatment procedures performed, prescriptions ordered, and billing charges on all outpatient and in-hospital encounters, with diagnoses and procedures coded based on the International Classification of Diseases, Ninth and Tenth revisions (ICD-9 and ICD-10, respectively).

We linked the deidentified primary data sources using common variables within the UPMC data systems aggregated in its Clinical Data Warehouse that include: (i) *Medipac*, the admit, discharge and transfer registration and hospital-based billing system; (ii) *Cerner*, the inpatient EMR for relevant clinical information for bedded patients at a UPMC inpatient hospital; (iii) *Epic*, the UPMC EMR for ambulatory office visits owned by UPMC; and (iv) *Aria*, the EMR utilized in most ambulatory Cancer Centers at UPMC for both radiation oncology and medical oncology.^8^

### Study Population

The study population was derived from patients who received bamlanivimab from December 9, 2020 to March 3, 2021. Patients were candidates for therapy based on criteria consistent with EUA criteria including: recently diagnosed mild to moderate COVID-19 (with a positive polymerase chain reaction or antigen test for SARS-CoV-2 virus within 10 days of the date the test was obtained); body mass at least 40 kg; age ≥65 years or age ≥12 years with a medical condition conferring high risk of COVID-19 progression to severe disease and/or hospitalization.^9^ Patients were included in the study population if they completed the mAb infusion and had attainment of one of the study outcomes of interest within 28 days of infusion, or had 28 days of follow-up without an outcome.

We also derived a comparator group from the same at-risk population by identifying non-hospitalized patients with a positive polymerase chain reaction or antigen test for SARS-CoV-2 during the same time period who were eligible for mAb infusion based on our modified EUA criteria but not infused. Similar to the group receiving mAb, comparator patients were included in the study population if they had attainment of one of the study outcomes of interest within 28 days or had 28 days of follow-up without an outcome. For infused patients, the 28-day follow-up period commenced on the date of their infusion. For comparator patients, the follow-up period commenced two days after their SARS-CoV-2 test result date, which corresponded to the earliest time from test positivity to initiation of treatment for infused patients.

### Study Outcomes

The primary outcome was hospitalization or all-cause mortality within 28 days of meeting study eligibility (day of infusion for the group receiving mAb, 30 days after test positivity for the non-infused group). We assessed in-hospital mortality using the discharge disposition of “Ceased to Breathe” sourced from the inpatient EMR and out-of-hospital deaths from the Social Security Death Index. The secondary outcome was hospitalization or 28-day Emergency Department (ED) visit without hospitalization. To understand the contribution of individual elements of the primary and secondary composite outcomes, we also analyzed the frequency of individual events: ED visit without hospitalization, hospitalization, and mortality within 28 days.

### Statistical Methods

After identifying patients in the study population receiving mAb, we selected from the at-risk comparator population individuals matched by propensity score.^10, 11^ Age is a strong predictor for complications of COVID-19 and the prevalence of high-risk medical conditions vary substantially by patient age.^12, 13^ We anticipated that differences in patient profiles between infused and non-infused patients would vary by age because of the age-specific criteria for bamlanivimab contained in the EUA. Moreover, immunosenescence occurs with aging, so we anticipated age to be a potential effect modifier in the relationship between mAb receipt and study outcomes.^14^ Therefore, we planned *a priori* to select non-infused patients using propensity scores within age strata consistent with the EUA criteria: less than 55 years, 55 years to less than 65 years, and 65 years of age and older.^9^ Patients receiving mAb infusion and the selected comparator patients across all age strata were then combined to constitute the study population. We used this approach, rather than initially running a single model from the full dataset, to optimize selection of appropriate controls. To illustrate, in the full dataset, the prevalence of history of morbid obesity was 27.4% versus 24.9% (p=0.37) in infused vs. non-infused patients, respectively, whereas in the subset of patients age 55 to < 65 years, the respective prevalence was 66.7% vs. 22.4% (p<0.00001). Thus, we believe that more robust matching of infused to non-infused patients was achieved by capturing and adjusting for the important differences in presenting profiles within each age-specific cohort.

Propensity scores were derived using logistic regression models fit from a multitude of variables in separate age-stratified groups with treatment with mAb as the response variable and forward stepwise selection of measured pre-treatment explanatory variables at p <0.15 (see **Supplemental Table 1** for listing of variables used in the propensity score models). We included variables deemed biologically relevant (e.g. age, gender, Charlson Comorbidity Index score) into all models prior to stepwise selection. We used 1:1 propensity score matching with a maximum propensity score probability difference of 0.01 to construct equal size matched infused and non-infused groups within age strata. We did not impute missing values for variables used in deriving the propensity scores but did assess whether patients with propensity scores (i.e., full covariate data) versus those with missing propensity scores were similar in terms of age, gender, and race distributions (to assess randomness of missing data).

We compared characteristics of infused versus non-infused patients using student *t*-tests for continuous variables and chi-square tests for categorical variables. Because in this observational study we must infer the time at risk for non-infused patients (starting at the time they *would have* received mAb had they been referred for care, conservatively using two days after SARS-CoV-2 testing), we compared the distribution of time from beginning of follow-up to study outcome among patients receiving mAb infusion and those who did not, both in the at-risk and matched populations (**Supplemental Table 2**). Crude 28-day rates of the study outcomes were described by infusion status. For the propensity matched patient analysis of primary and secondary outcomes, as well as individual elements of the composite outcomes (ED visit without hospitalization, hospitalization, mortality) we fit logistic regression models with the derived propensity scores (i.e., predicted probability of being treated with mAb) as a continuous variable to control for confounding, with mAb receipt as the predictor of interest in the model.

We also performed one sensitivity and three exploratory analyses. As a sensitivity analysis, we used the propensity score (i.e., predicted probability of being treated with mAb) as a continuous variable to control for confounding and evaluate study outcomes in a larger unmatched cohort of patients who did and did not receive mAb. We did not employ the use of inverse probability weighting given the very large imbalance of eligible bamlanivimab treated versus not infused patients. We postulated that mAb may be more efficacious in the potentially immunosenescent older population. Thus, we evaluated the study outcomes within the pre-defined age strata. In the second exploratory analysis among the larger unmatched cohort, an interaction term of mAb x BMI was created to examine potential effect modification of treatment with mAb by BMI.^15^ To identify a potential benefit of prompt (versus delayed) administration of mAb, as a third exploratory analysis we compared the rates of three outcomes – ED visit without hospitalization, hospitalization, and mortality – within the analysis group stratified by time from diagnosis to infusion: 0 to 2 days, 3 to 4 days, 5 to 7 days, and 8 to 10 days.

Study outcomes are described with effect estimate (odds ratio [OR]) and 95% confidence interval (95%CI). We set the alpha error at 0.05 for univariate comparisons of baseline characteristics by infusion status as well as for adjusted odds ratios. All analyses were performed using the SAS System (Cary, NC), version 9.4. Methods and results are reported in accordance with The REporting of studies Conducted using Observational Routinely-collected health Data (RECORD) statement (see **Supplemental Table 3**).^16^

## RESULTS

### Study Population

During the study period (December 9, 2020 through March 3, 2021), 636 patients received mAb. Four hundred and sixty-three (72.8%) patients achieved a study outcome or completed 28-day follow-up time and are in the analysis group, with the last infused patient who did not die treated on February 3, 2021. The non-infused at-risk population included 17,599 COVID-19 non-hospitalized patients potentially eligible for mAb according to modified EUA criteria, of which 16,565 (94.1%) achieved study outcome or had sufficient follow-up time.

**Table 1** shows a comparison of baseline characteristics of the infused versus non-infused patients both in the unmatched at-risk population and matched study population. The unmatched populations are significantly different on most characteristics with patients receiving mAb infusion demonstrating higher frequencies of these medical conditions. After propensity score matching within age strata, 234 patients receiving mAb infusion and 234 patients not receiving mAb were included in the study population. After propensity matching, all matched and selected unmatched variables did not differ statistically between patients who received versus did not receive mAb infusion.

**Table 1.**
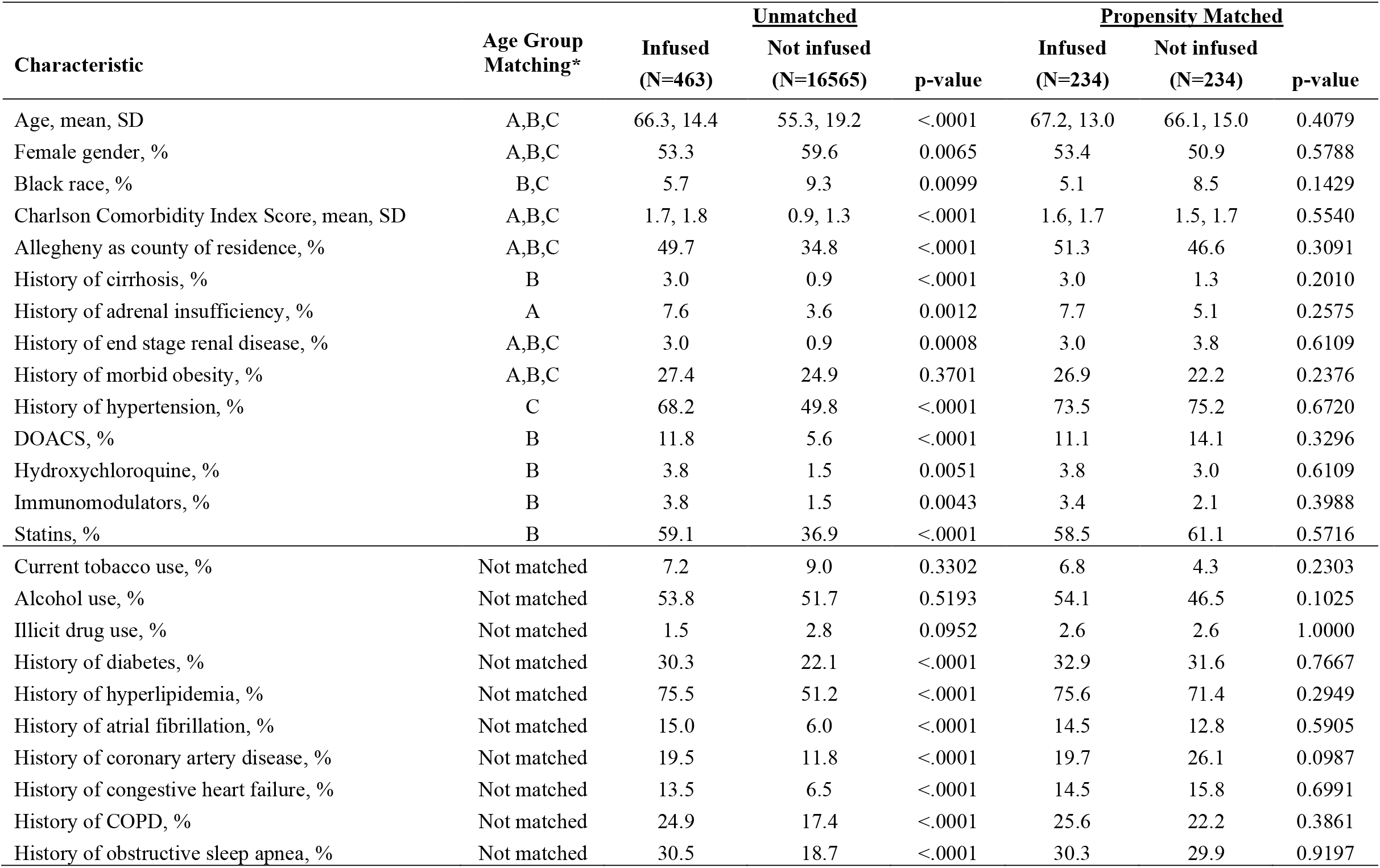
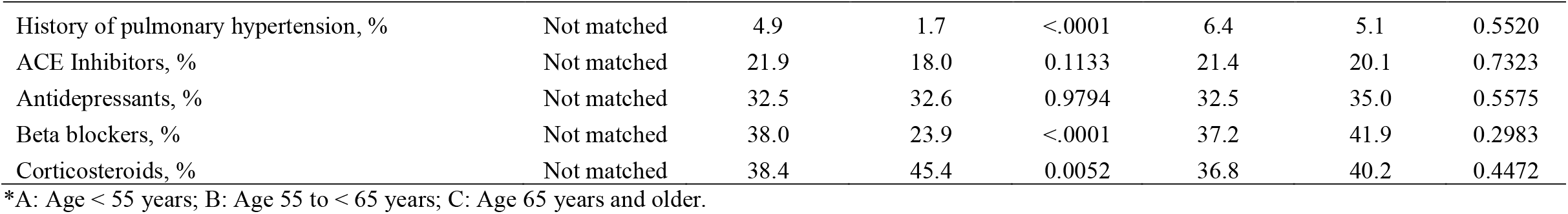
Comparison of Characteristics of Unmatched and Propensity Matched Patients.

### Primary and Secondary Outcomes

Among the 234 propensity-matched patients receiving mAb infusion, 15 (6.4%) were hospitalized, 4 (1.7%) died, and 16 (6.8%) had an ED visit without hospitalization (**Table 2**). Among the 234 propensity-matched patients not receiving mAb infusion, 38 (16.2%) were hospitalized, 12 (5.1%) died, and 15 (6.4%) had an ED visit without hospitalization. **Figure 1a** shows the frequency of the primary outcome (28-day hospitalization or mortality) and the secondary outcome (hospitalization or ED visit without hospitalization) among the matched patients receiving mAb versus those not receiving mAb infusion. In the propensity adjusted analyses (**Table 2**), patients receiving mAb had an estimated 69% lower odds of hospitalization or mortality (OR 0.31, 95% CI 0.17-0.56) and estimated 50% lower odds of hospitalization or ED visit without hospitalization (OR 0.50, 95% CI 0.43-0.83). **Figure 1b** shows the frequency of the individual elements of the composite primary and secondary outcomes (ED visit without hospitalization, hospitalization, mortality) among the matched patients receiving mAb versus those not receiving mAb infusion, and **Table 2** lists the corresponding ORs and 95% confidence intervals.

**Table 2.** Propensity Matched Event Rates and Odds Ratios of Study Outcomes Overall and Stratified by Age Group.

| Outcome | Number of Events |  | 28-Day Event Rate (%) |  | Odds Ratio Estimates |  |  |
| --- | --- | --- | --- | --- | --- | --- | --- |
|  | Infused<br>(n=234) | Not Infused<br>(n=234) | Infused | Not Infused | Odds<br>Ratio | 95% CI | p-value |
| **All Patients** |  |  |  |  |  |  |  |
| Hospitalization or mortality | 16 | 45 | 6.8 | 19.2 | 0.31 | 0.17, 0.56 | 0.00001 |
| Hospitalization or ED visit without hospitalization | 28 | 50 | 12.0 | 21.4 | 0.50 | 0.43, 0.83 | 0.007 |
| ED visit without hospitalization | 16 | 15 | 6.4 | 6.4 | 1.07 | 0.52, 2.22 | 0.85 |
| Hospitalization | 15 | 38 | 6.4 | 16.2 | 0.35 | 0.19, 0.66 | 0.001 |
| Mortality | 4 | 12 | 1.7 | 5.1 | 0.32 | 0.10, 1.01 | 0.05 |
| **Age < 55 years** | **(n=42)** | **(n=42)** |  |  |  |  |  |
| Hospitalization or mortality | 2 | 3 | 4.8 | 7.1 | 0.65 | 0.10, 4.10 | 0.65 |
| Hospitalization or ED visit without hospitalization | 7 | 5 | 16.7 | 11.9 | 1.48 | 0.43, 5.10 | 0.53 |
| ED visit without hospitalization | 6 | 4 | 14.3 | 9.5 | 1.58 | 0.41, 6.08 | 0.50 |
| Hospitalization | 2 | 3 | 4.8 | 7.1 | 0.65 | 0.10, 4.10 | 0.65 |
| Mortality | 0 | 0 | 0.0 | 0.0 | ----- | ----- | ----- |
| **Age 55 to < 65 years** | **(n=34)** | **(n=34)** |  |  |  |  |  |
| Hospitalization or mortality | 2 | 6 | 5.9 | 17.6 | 0.29 | 0.05, 1.56 | 0.15 |
| Hospitalization or ED visit without hospitalization | 2 | 7 | 5.9 | 20.6 | 0.24 | 0.05, 1.26 | 0.09 |
| ED visit without hospitalization | 1 | 2 | 2.9 | 5.9 | 0.49 | 0.04, 5.61 | 0.56 |
| Hospitalization | 2 | 5 | 5.9 | 14.7 | 0.36 | 0.06, 2.01 | 0.25 |
| Mortality | 0 | 1 | 0.0 | 2.9 | 0.0 | ----- | ----- |
| **Age 65 years and older** | **(n=158)** | **(n=158)** |  |  |  |  |  |
| Hospitalization or mortality | 12 | 36 | 7.6 | 22.8 | 0.28 | 0.14, 0.56 | 0.0003 |
| Hospitalization or ED visit without hospitalization | 19 | 38 | 12.0 | 24.0 | 0.43 | 0.24, 0.79 | 0.006 |
| ED visit without hospitalization | 9 | 9 | 5.7 | 5.7 | 1.00 | 0.39, 2.59 | 1.0 |
| Hospitalization | 11 | 30 | 7.0 | 19.0 | 0.32 | 0.15, 0.66 | 0.002 |
| Mortality | 4 | 11 | 2.5 | 7.0 | 0.35 | 0.11, 1.11 | 0.08 |

**Figure 1a.**
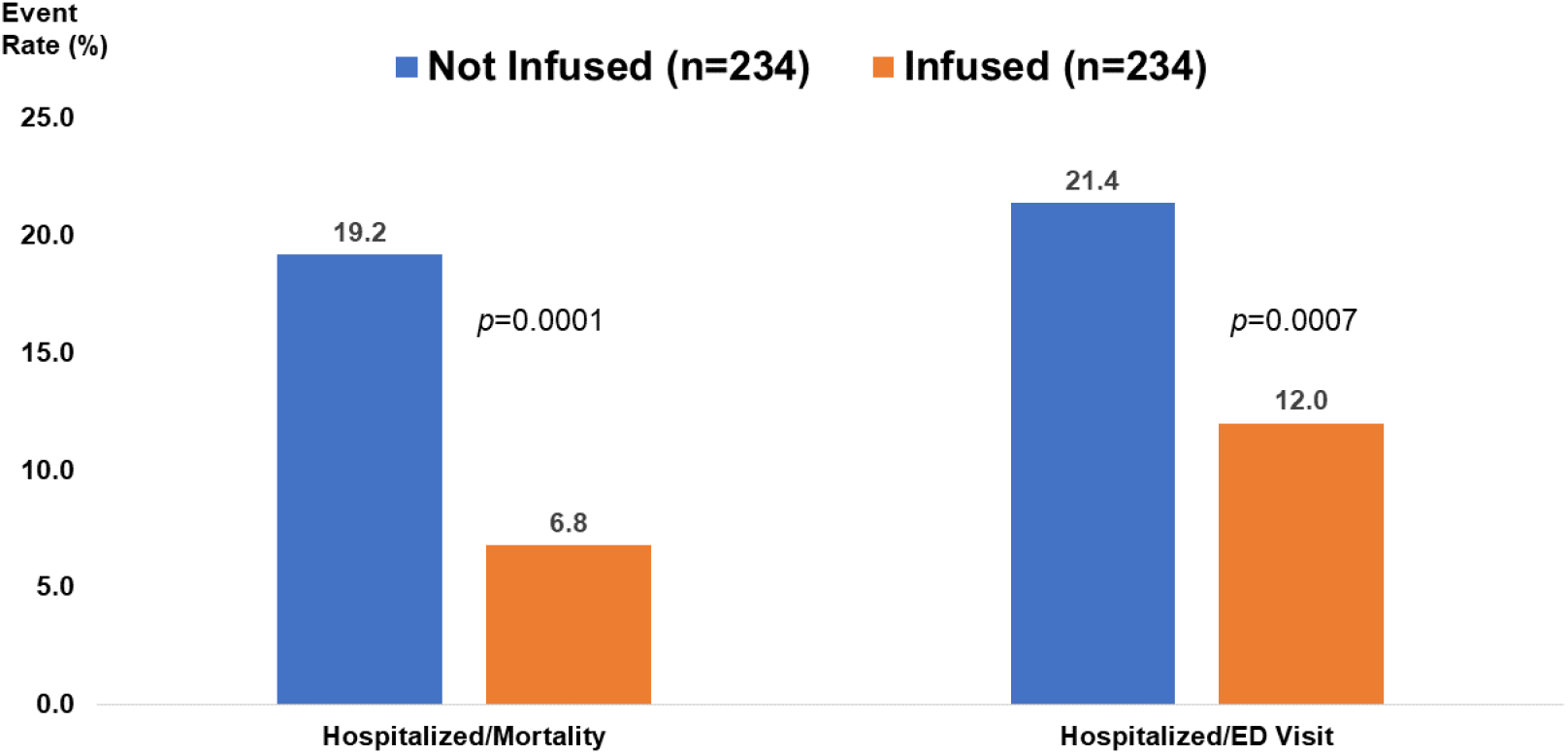
Frequency of 28-day hospitalization or mortality (primary outcome), and hospitalization or Emergency Department visit without hospitalization (secondary outcome) among the matched patients receiving monoclonal antibody (orange bars) versus those not receiving monoclonal antibody infusion (blue bars). P-values are from the logistic regression models.

**Figure 1b.**
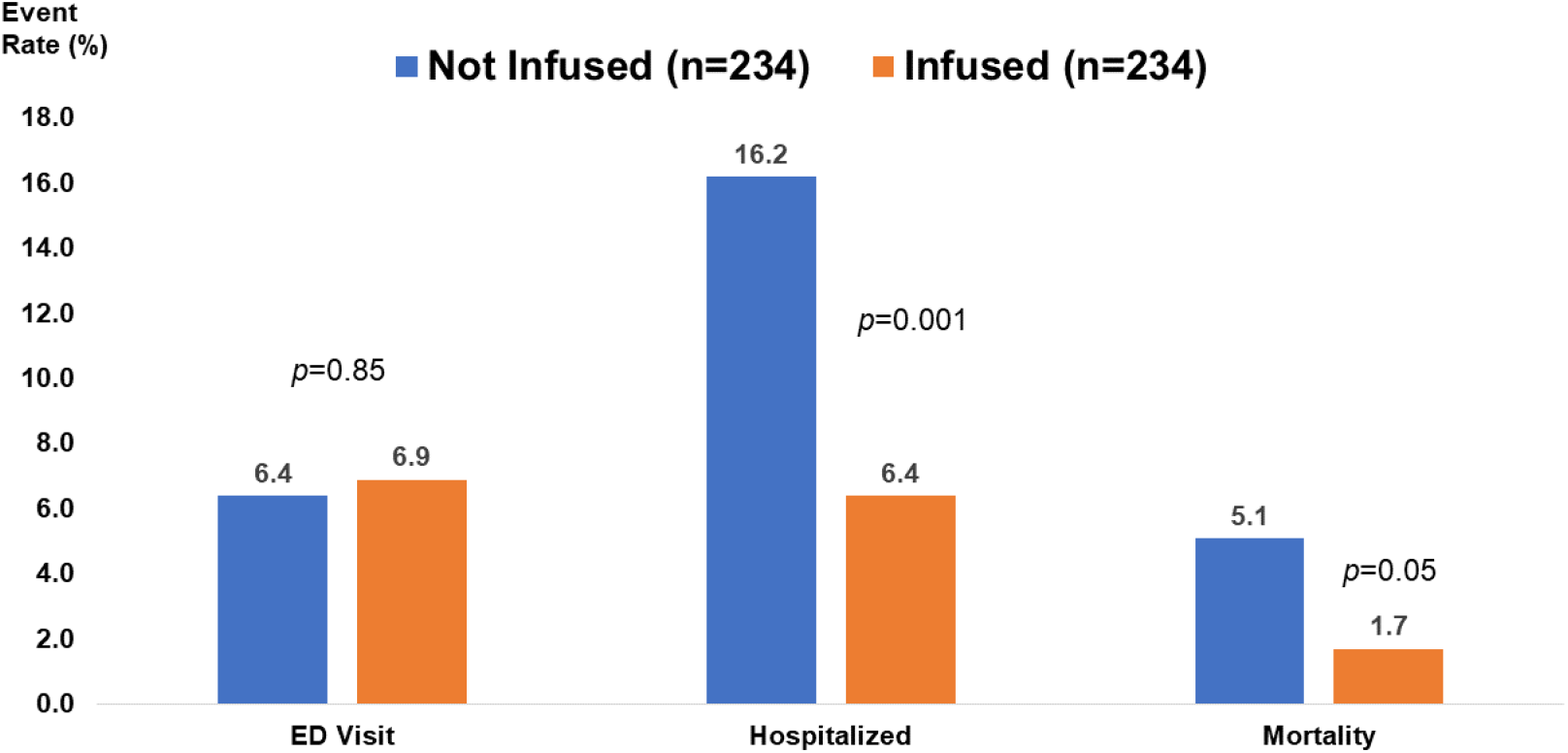
Frequency of the individual elements of the composite primary and secondary outcomes including Emergency Department visit without hospitalization, hospitalization, and mortality among the matched patients receiving monoclonal antibody (orange bars) versus those not receiving monoclonal antibody infusion (blue bars). P-values are from the logistic regression models.

### Sensitivity and Exploratory Analyses

**Supplemental Table 4** provides risk estimates for the composite primary and secondary outcomes, and outcomes comprising the composite outcomes, in an unmatched cohort of patients receiving monoclonal antibody infusion and an at-risk population of patients not receiving monoclonal antibody infusion, adjusted for propensity to receive mAb. The 236 patients with non-missing covariate data who received mAb had an estimated 59% lower odds of hospitalization or mortality than the 16,565 patients who did not receive mAb (propensity-score adjusted OR 0.41, 95% CI 0.24-0.70). Thus, study results were consistent between the matched and unmatched group analyses.

**Table 2** provides adjusted ORs for the primary and secondary outcomes, plus individual outcomes comprising the composite outcomes, stratified by age. In propensity score adjusted analyses for the 28-day rate of hospitalization or mortality, there was no differential treatment effect by BMI. P-values for the interaction term of mAb x BMI were 0.55, 0.81, 0.38, and 0.84 for all patients and in the age groups less than 55 years, 55 to less than 65 years, and 65 years and older, respectively. **Supplemental Figure 1** indicates among patients aged 65 years and older lower composite hospitalization or mortality event rates comparing mAb infused to non-infused patients across all BMI categories. **Figure 2** indicates that among patients in the study population receiving mAb infusion, those who received their infusion within 4 days of their positive SARS-CoV-2 test result had lower 28-day rates of ED visit without hospitalization and hospitalization than patients who received their infusion 5 days or more after their positive SARS-CoV-2 test result. Rates of mortality were low and similar by timing of infusion.

**Figure 2.**
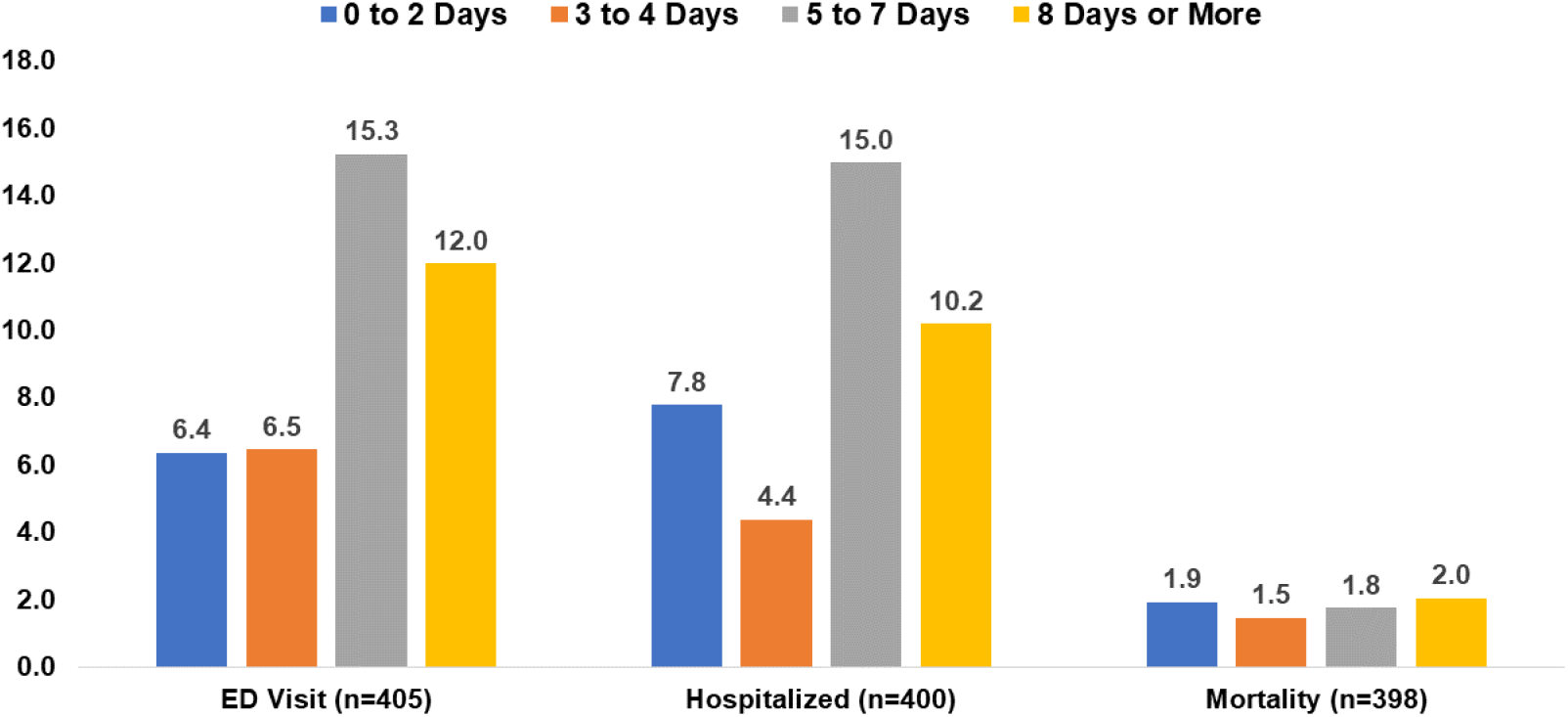
Among all study patients receiving monoclonal antibody infusion, comparison of crude 28-day outcome rates by timing of infusion. Blue bars (0 to 2 days), orange bars (3 to 4 days), grey bars (5 to 7 days), yellow bars (8 days or more).

Finally, age, gender, and race distributions between patients with full covariate data and those with missing data were similar (data not shown).

## DISCUSSION

Our observational study suggests a benefit of mAb treatment in reducing the risk of COVID-19 complications for patients at higher risk of severe disease. Initial reports of bamlanivimab 700 mg in outpatients showed improvement of symptoms at day 11 for the entire study population^3^ and numerically fewer hospitalizations and ED visits at day 29 (1.6% vs 6.3%).^17^ A post hoc analysis in BLAZE-1 of those 65 or older or with BMI of 35 kg/m^2^ or greater demonstrated a larger numerical benefit, 2.7% vs 13.5%.^3^ In this report of our experience, we identified a similar association between bamlanvimab monotherapy and hospitalization or mortality in a propensity matched analysis with an OR of 0.31, or a 69% lower odds of hospitalization or mortality.

In our health system, rates of hospitalizations and ED visits in patients aged 65 years or older were higher than the clinical trial, yet a benefit to mAb treatment was nonetheless observed. This benefit occurred through decreased hospitalizations and mortality (OR 0.28) as well as for hospitalizations alone (OR 0.32) and for hospitalizations or ED visits without hospitalization (OR 0.43). We also identified a tendency of mortality benefit in this age group (OR 0.35), although our sample size and event limit declaring an association.

Cumulatively our report shows benefit for patients across several possible outcomes, and we consider this highly relevant from a clinical standpoint. While we did not observe statistically significant differences in the primary or secondary outcomes in younger age groups, the odds ratios in the group aged 55 to less than 65 years, and few outcomes in those age less than 55 years suggest an effect may exist though potentially smaller in magnitude.

We identified greater benefits with administration within 4 days of symptom onset, particularly regarding rates of hospitalization. While the EUA allows for use within 10 days of symptom onset, the median duration of symptoms prior to receiving bamlanivimab 700 mg was 5 days in BLAZE-1^3^. As with other passive antibody therapies, this is consistent with the supposition that earlier treatment is better, and health systems may reasonably evaluate a shorter window of eligibility for mAb therapy. Our analysis shows that benefits in the elderly are independent of BMI. The potential differential benefit of mAb therapy for COVID-19 in patients with this and other co-morbidities warrants more focused analysis.

There are several limitations of our study. Given our design, it is not surprising that there were several baseline differences between those infused and not infused in other age groups. We mitigated confounding with propensity score modeling of closely matched groups of patients, and a sensitivity analysis using propensity-score adjusted unmatched patients not receiving mAb infusion affirmed the findings. Furthermore, the group receiving mAb had more comorbid conditions predisposing to the primary and secondary outcomes, which may underestimate the magnitude of the treatment effect if there was residual confounding. We cannot reliably distinguish the presence, extent, or severity of symptoms in our data set, all of which may impact effectiveness. Additionally, viral loads in blood or any site were not measured in our data set, limiting any insights based on this variable. The time to event in both treated and untreated groups were similar, suggesting that ED visit or hospitalization did not account for why the untreated population did not receive mAb. Viral loads in blood or any site were not measured in our data set, limiting any insights based on this variable.

We did not have information regarding variant strains of SARS-CoV-2 prior to or after bamlanivimab monotherapy. While reported rates of clinically concerning variants were low in Pennsylvania during much of this time frame, there is concern that use of bamlanivimab monotherapy will lead to escape variants and/or that variants are underreported.^18^ We were unable to evaluate any such existing prevalence or emergence in our patients. Finally, during the time of this study, we utilized bamlanivimab monotherapy exclusively, so we are unable to comment on any comparison to the other available monoclonal antibodies for treatment of mild-moderate COVID-19 infection. On March 24^th^, the United States Department of Health and Human Services announced they would no longer supply sites with bamlanivimab alone due to concern about increased rates of resistant variants.^19^ Many of our patients received bamlanvimab when the rates of resistant variants in this country was low, thus explaining why we still saw benefit with bamlanivimab monotherapy. Going forward it will be critical to better define the roles of the various available therapies and where each may be best utilized, including possibly a continued role for bamlanivimab monotherapy.

In our non-experimental design, we observed that bamlanivimab monotherapy is associated with decreased hospitalization rates and mortality in patients with mild-moderate COVID-19 infection, particularly among those 65 years or older, and may likely extend to younger patients as well. This benefit appears more likely when administered early after diagnosis. Further study can confirm our observations and investigate the role of mAb treatment in other high-risk subgroups and the use of various mAb regimens.

## Data Availability

The data was used from electronic medical records as part of quality improvement and is not publicly available.

## Acknowledgements

The authors thank the clinical staff of the UPMC monoclonal antibody infusion centers as well as the support and administrative staff behind this effort, including but not limited to: Debbie Albin, Jennifer Dueweke, Robert Shulik, Amy Lukanski, Rozalyn Russell, Debra Rogers, Jesse Duff, Kevin Pruznak, Jennifer Zabala, Trudy Bloomquist, Daniel Gessel, LuAnn King, Jonya Brooks, Libby Shumaker, Betsy Tedesco, Sarah Sakaluk, Kathleen Flinn, Susan Spencer, Le Ann Kaltenbaugh, Michelle Adam, Meredith Axe, Melanie Pierce, Debra Masser, Theresa Murillo, Sherry Casali, Jim Krosse, Jeana Colella, Rebecca Medva, Jessica Fesz, Ashley Beyerl, Jodi Ayers, Hilary Maskiewicz, Mikaela Bortot, Amy Helmuth, Heather Schaeffer, Janice Dunsavage, Erik Hernandez, Ken Trimmer, Sheila Kruman, Teressa Polcha, and their entire teams. We also thank the U.S. federal government and Pennsylvania Department of Health for the provision of monoclonal antibody treatment.

## SUPPLEMENTAL MATERIALS

**Supplemental Table 1.** Variables selected for propensity score adjustment

| Analysis Group | Variables |
| --- | --- |
| Age < 55 years | Age<br>Gender<br>Residence in Allegheny county<br>Morbid obesity<br>History of renal insufficiency<br>History of end stage renal disease<br>History of irritable bowel syndrome<br>Charlson Comorbidity Index score<br>Model c-statistic: 0.826 |
| Age 55 to < 65 years | Age<br>Gender<br>Race<br>Residence in Allegheny county<br>Morbid obesity<br>History of cirrhosis<br>History of end stage renal disease<br>Charlson Comorbidity Index score<br>Medications: direct oral anticoagulants, hydroxychloroquine, immunomodulators, statins<br>Model c-statistic: 0.852 |
| Age ≥ 65 years | Age<br>Gender<br>Race<br>Residence in Allegheny county<br>Morbid obesity<br>History of hypertension<br>Charlson Comorbidity Index score<br>Model c-statistic: 0.670 |

**Supplemental Table 2.** Comparison of days to individual outcomes between patients receiving monoclonal antibody infusion and patients not receiving monoclonal antibody infusion, in both the at-risk population and matched study population.

|  | Outcome | Infused |  |  | Not Infused |  |  | p-value |
| --- | --- | --- | --- | --- | --- | --- | --- | --- |
|  |  | N | Median | IQR | N | Median | IQR |  |
| At-risk population | ED visit without hospitalization | 30 | 7 | 4, 15 | 1040 | 4 | 2, 10 | 0.04 |
|  | Hospitalization | 31 | 8 | 2, 15 | 1379 | 5 | 2, 11 | 0.40 |
|  | Mortality | 7 | 13 | 5, 23 | 301 | 11 | 7, 16 | 0.38 |
| Matched study population | ED visit without hospitalization | 16 | 7 | 2.5, 14 | 15 | 4 | 2, 8 | 0.20 |
|  | Hospitalization | 15 | 5 | 1, 21 | 38 | 8.5 | 3, 16 | 0.66 |
|  | Mortality | 4 | 18 | 8.5, 24 | 12 | 9.5 | 9, 11 | 0.16 |

Note: ED, Emergency Department; IQR, interquartile range

**Supplemental Table 3.**
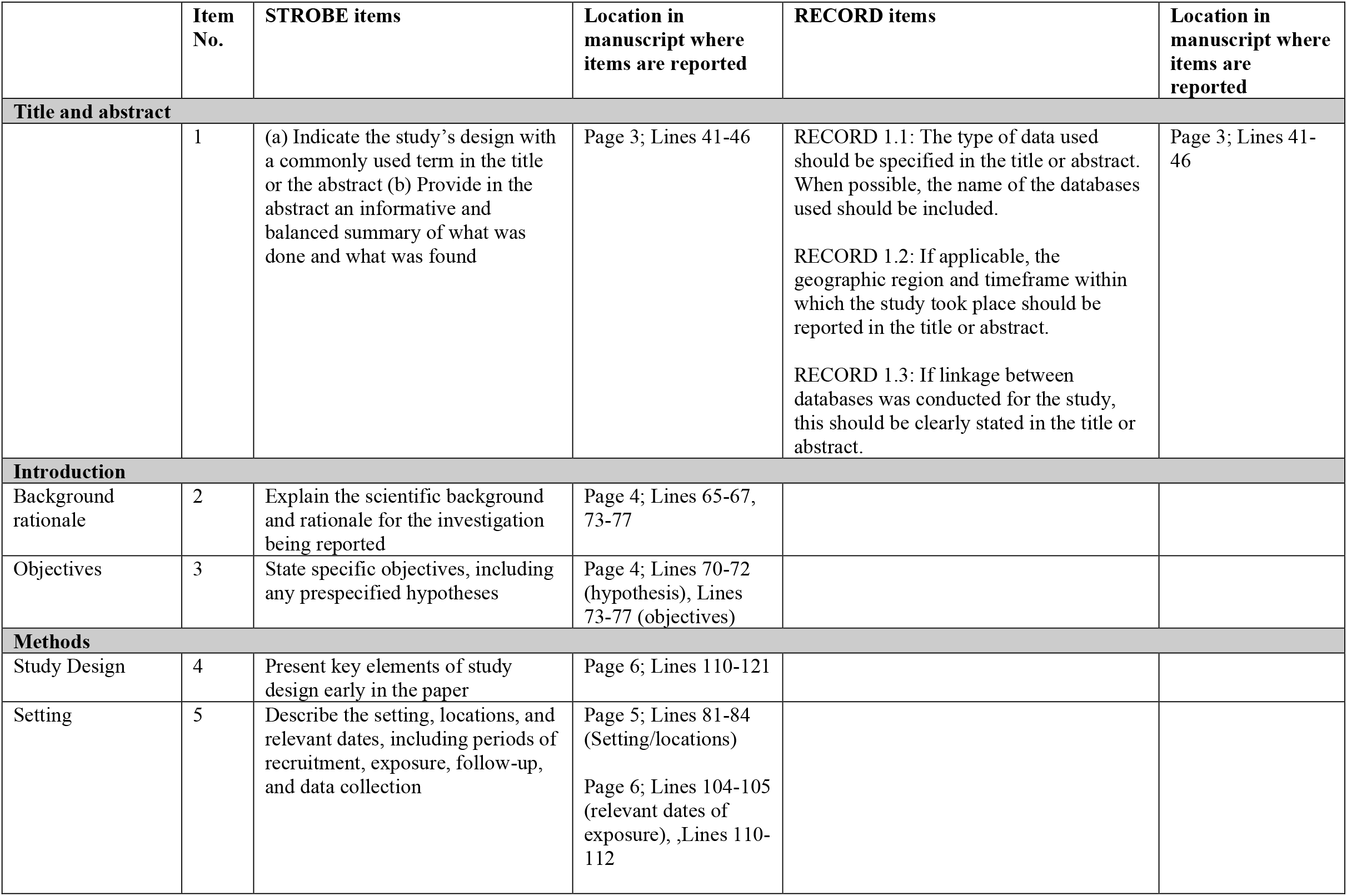
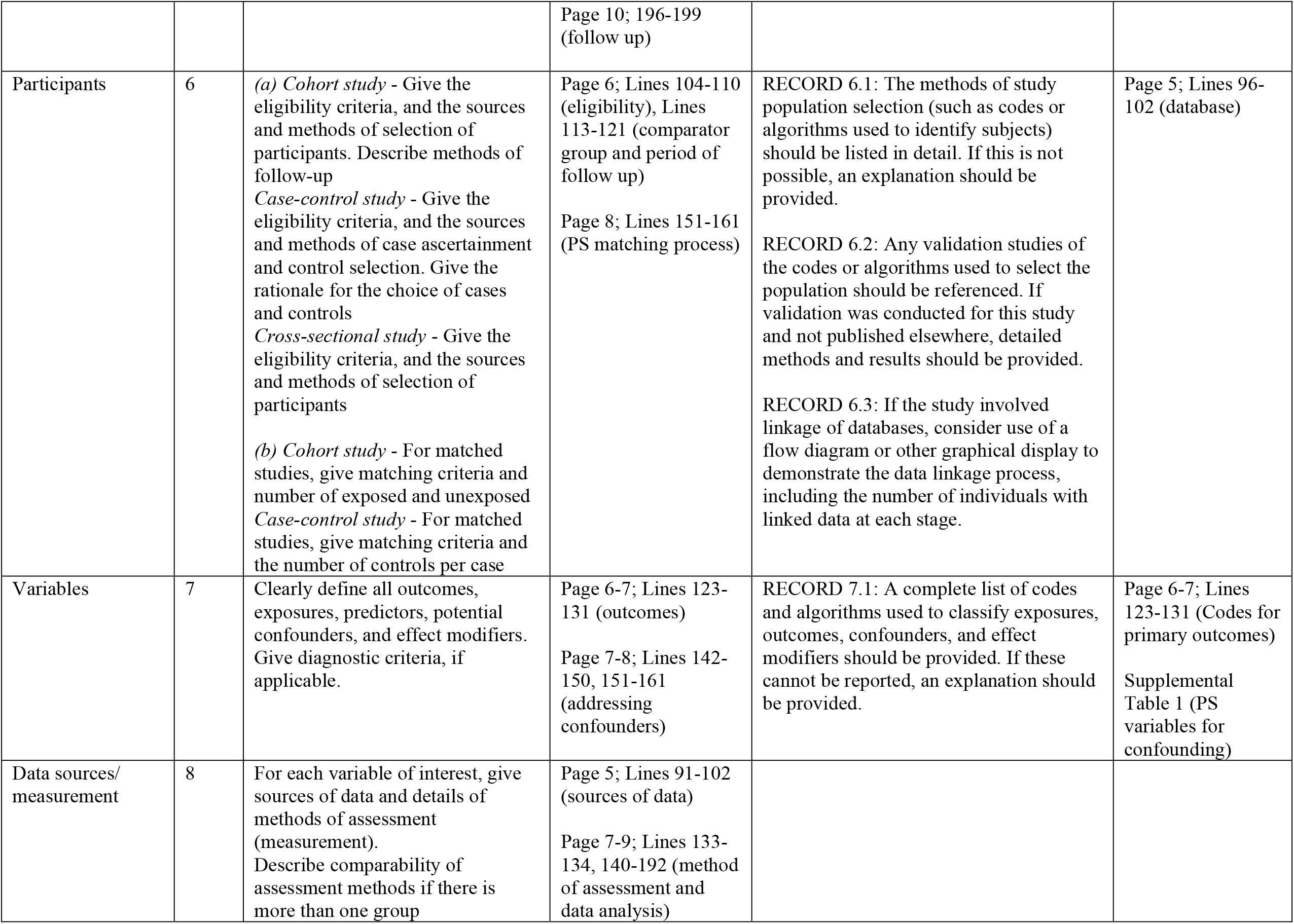
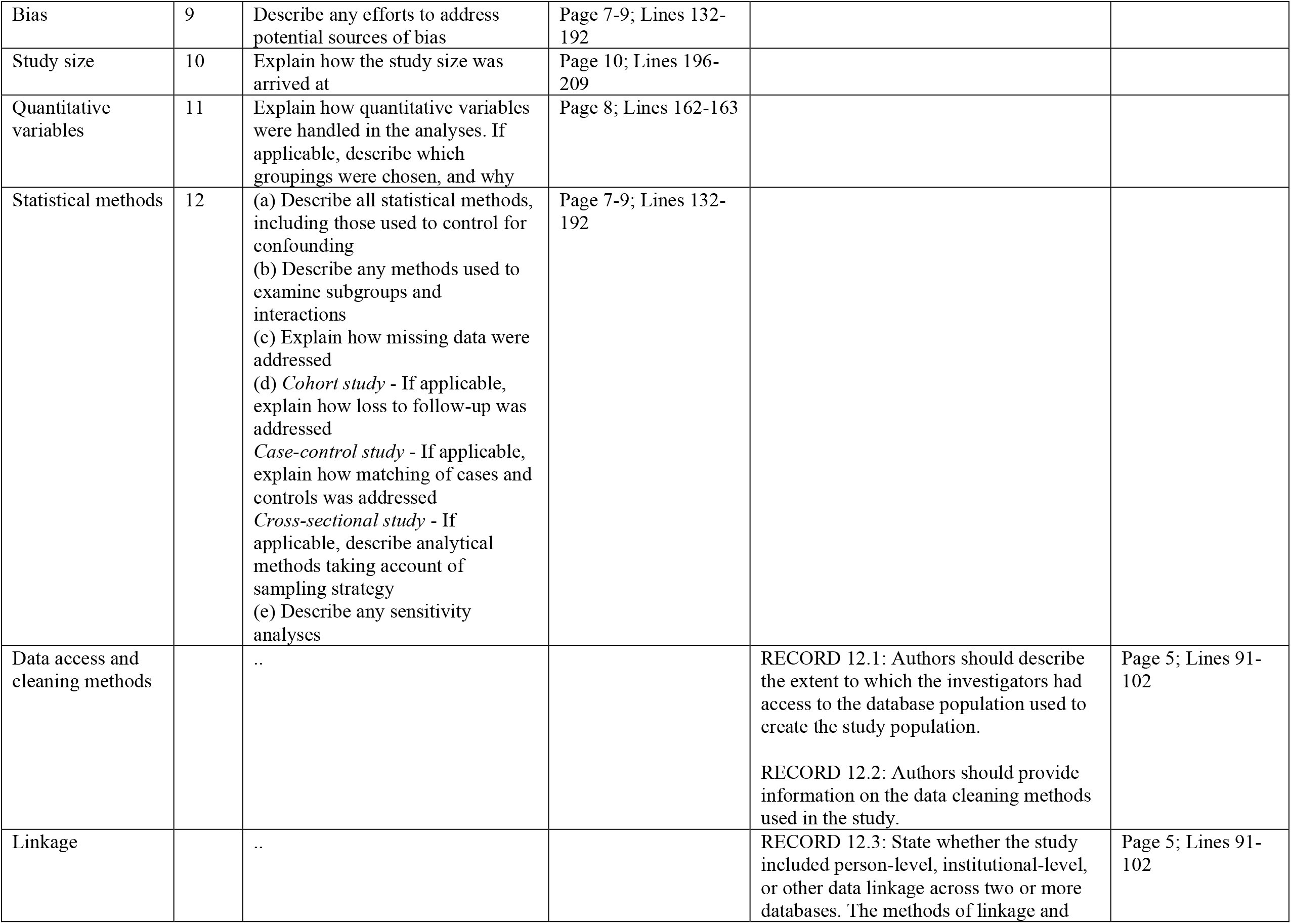
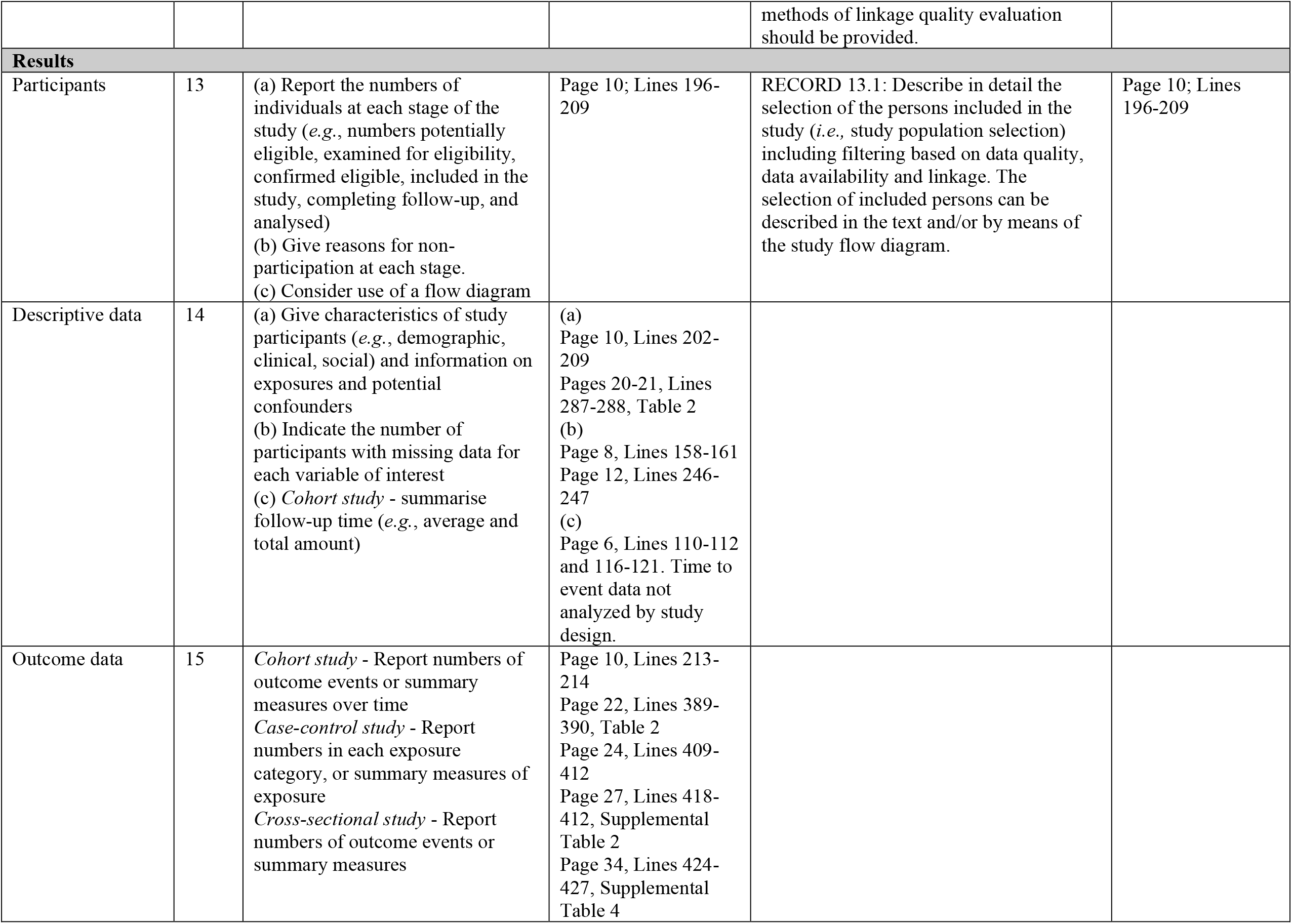
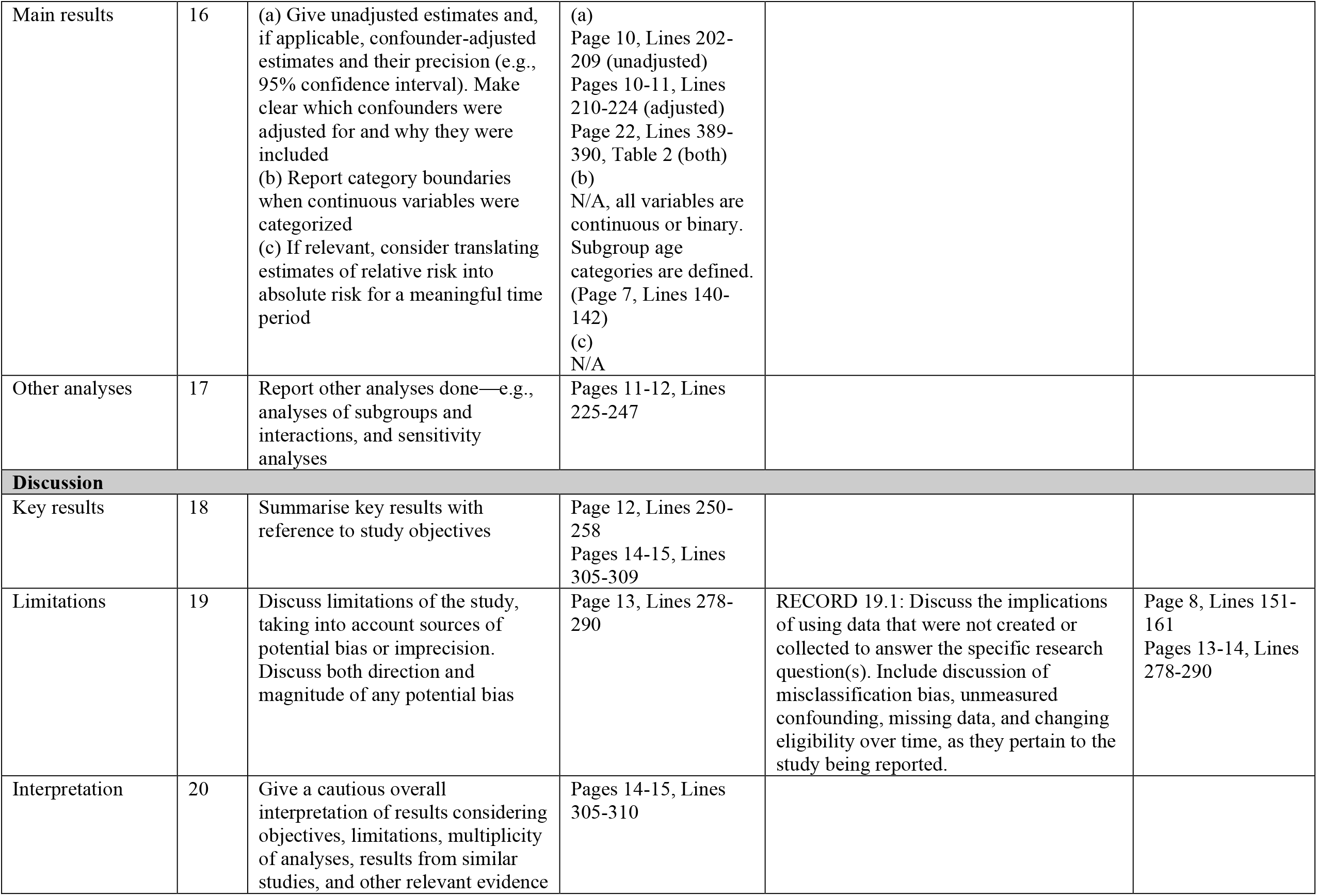
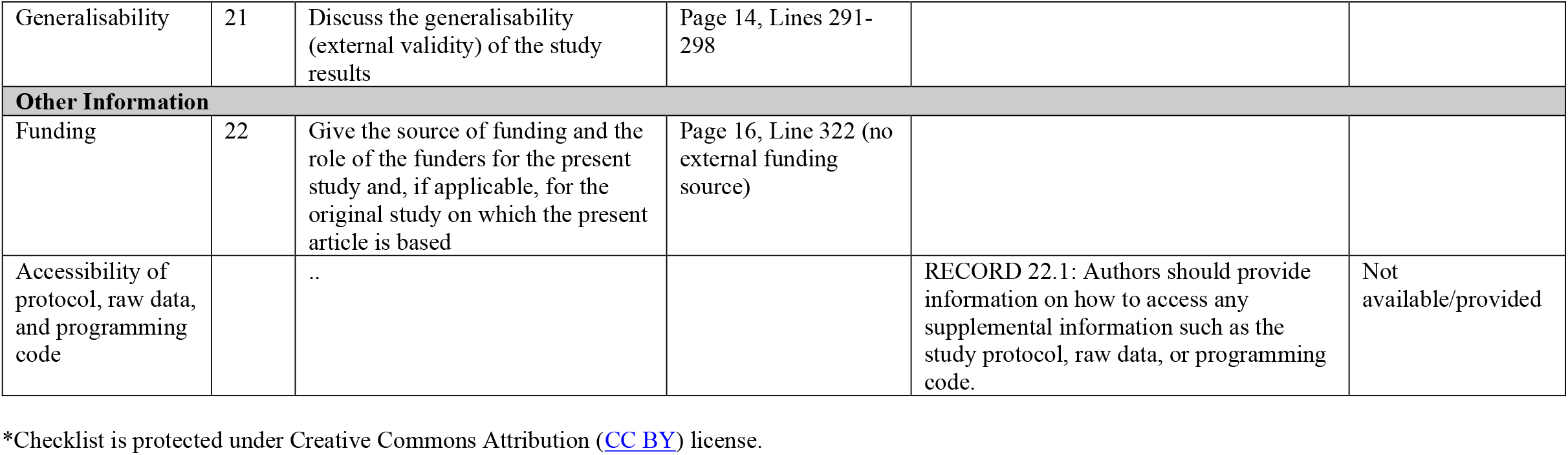
RECORD statement

**Supplemental Table 4.** Composite primary and secondary outcomes, and outcomes comprising the composite outcomes, in an unmatched cohort of patients receiving monoclonal antibody infusion and an at-risk population of patients not receiving monoclonal antibody infusion

|  | All Patients |  |  |  | Patients with Propensity Score |  |  |  | Odds Ratio Estimates |  |  |
| --- | --- | --- | --- | --- | --- | --- | --- | --- | --- | --- | --- |
|  | Number of Events |  | Event Rate (%) |  | Number of Events |  | Event Rate (%) |  |  |  |  |
|  | Infused<br>(n=463) | Not<br>Infused<br>(n=16,565) | Infused | Not<br>Infused | Infused<br>(n=236) | Not<br>Infused<br>(n=16,515) | Infused | Not<br>Infused | Adj. OR | 95%CI | P-value |
| Hospitalization or mortality | 33 | 1516 | 7.1 | 9.1 | 17 | 1507 | 7.2 | 9.1 | 0.41 | 0.24, 0.70 | 0.001 |
| Hospitalization or ED visit without hospitalization | 56 | 2254 | 12.1 | 13.6 | 29 | 2244 | 12.3 | 13.6 | 0.60 | 0.40, 0.91 | 0.02 |
| ED visit without hospitalization | 30 | 1040 | 6.5 | 6.3 | 16 | 1037 | 6.8 | 6.3 | 1.05 | 0.63, 1.77 | 0.85 |
| Hospitalization | 31 | 1379 | 6.7 | 8.3 | 16 | 1371 | 6.8 | 8.3 | 0.44 | 0.25, 0.77 | 0.004 |
| Mortality | 7 | 301 | 1.5 | 1.8 | 5 | 300 | 2.1 | 1.8 | 0.37 | 0.12, 1.16 | 0.29 |

Note: ED, Emergency Department; OR, odds ratio; 95%CI, 95% confidence interval

**Supplemental Figure 1.**
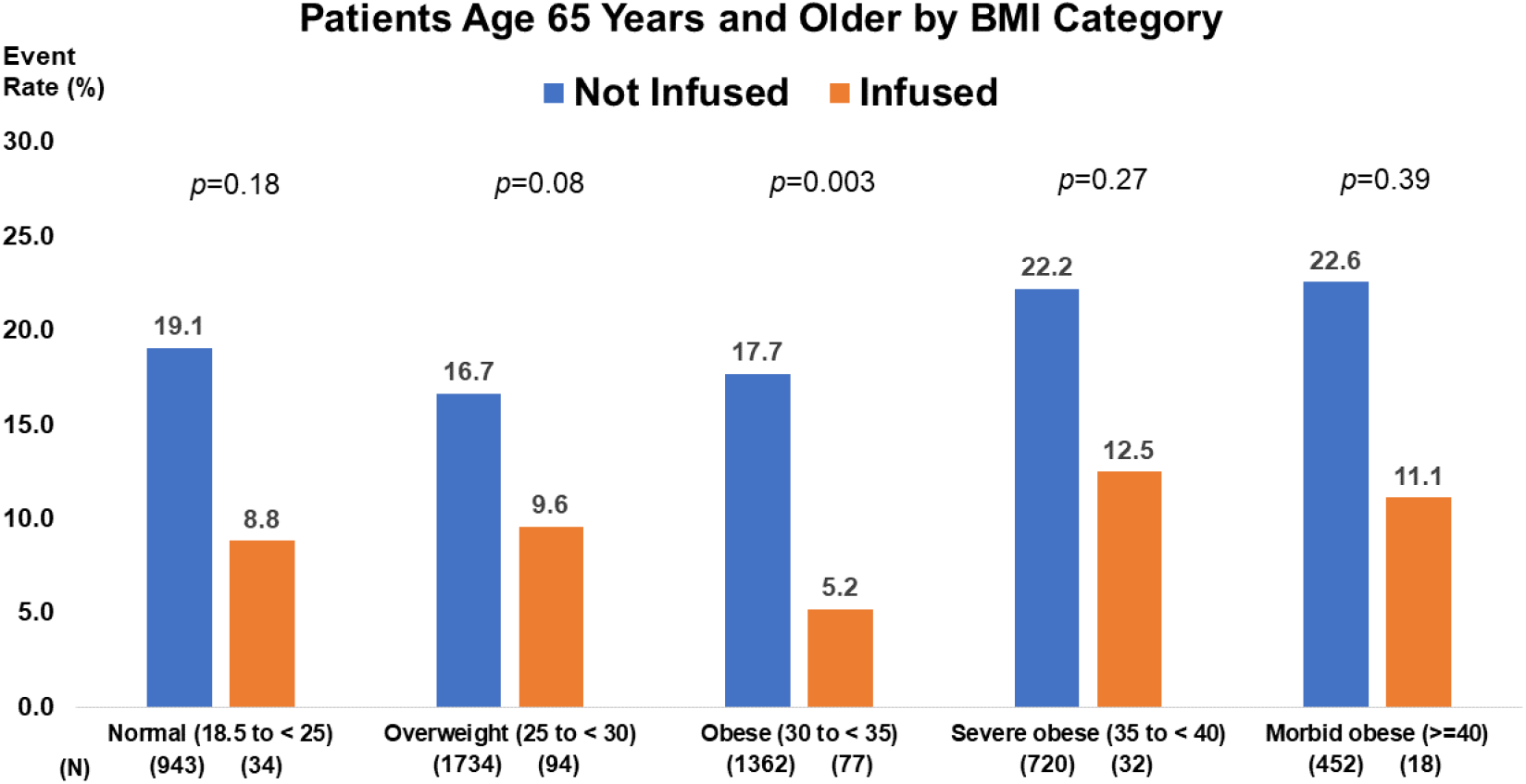
Frequency of composite hospitalization or mortality outcome among study population aged 65 years and older, by treatment received and BMI category Note: BMI, body mass index in units of kilograms per meter squared.

